# Complete remission in a patient with widely metastatic HER2-amplified pancreatic adenocarcinoma following multimodal therapy informed by tumor sequencing and organoid profiling

**DOI:** 10.1101/2021.12.16.21267326

**Authors:** Daniel A King, Amber R Smith, Gino Pineda, Michitaka Nakano, Flavia Michelini, S. Peter Goedegebuure, Sheeno Thyparambil, Wei-Li Liao, Aaron McCormick, Jihang Ju, Michele Cioffi, Xiuli Zhang, Jasreet Hundal, Malachi Griffith, Carla Grandori, Maddy Pollastro, Rachele Rosati, Astrid Margossian, Payel Chatterjee, Trevor Ainge, Marta Flory, Paolo Ocampo, Lee-may Chen, George A Poultsides, Ari D Baron, Daniel T Chang, Joseph M Herman, William E Gillanders, Haeseong Park, William A Hoos, Mike Nichols, George A Fisher, Calvin J Kuo

## Abstract

Here, we demonstrate a complete clinical response achieved in a patient with HER2+ metastatic pancreatic ductal adenocarcinoma to a coordinated barrage of anti-HER2, personalized vaccine and checkpoint inhibition immunotherapy, radiation, and chemotherapy. Comprehensive organoid profiling with drug sensitivity screening and drug testing suggested a vulnerability to anti-HER2 directed therapy, facilitating personalized treatment selection for our patient, which contributed to her clinical benefit. Immune response monitoring following personalized vaccine, radiation and checkpoint inhibition showed a sustained increase in neoantigen specific T cell response.

## Introduction

Pancreatic ductal adenocarcinoma (PDAC) remains among the deadliest of all human cancers^1^, compelling discovery of new predictive biomarkers and tailored therapies for this refractory disease. HER2 is a cell membrane receptor tyrosine kinase whose overexpression precipitates oncogenesis in several cancer types and is actively explored as a therapeutic target^2^.

HER2 overexpression in PDAC is uncommon, occurring in 2.1% of patients, of which 1.5% exhibit grade 3+ immunohistochemistry (IHC) positivity. The remaining 0.6% are IHC grade 2+ with amplification confirmed by *fluorescent in situ hybridization* (FISH)^3,4^. The role of HER2 as a prognostic and predictive biomarker in PDAC is controversial. While early studies indicated that HER2 amplification in PDAC may be a poor prognostic feature^5,6^, these findings were confounded by an overestimation of HER2 expression^3,7^. In contrast, both a recent large study^4^ and separate meta-analysis that measured HER2 amplification by FISH^8^, did not observe significant association between HER2 amplification and survival in PDAC. These data suggest that HER2 is not strongly prognostic in PDAC. Regarding HER2 as a biomarker of response to anti-HER2 directed therapy in PDAC, in vitro and in vivo animal models have indicated dose-dependent and HER2-expression-correlated survival improvements^9–12^. However, to date, human trials evaluating trastuzumab in combination with gemcitabine^13^, capecitabine^14^, or gemcitabine and erlotinib^15^, have shown median overall survival of 6.9-7.9 months, consistent with lackluster clinical benefit compared to standard of care therapy. Notably, these HER2-unselected PDAC trials measured a higher frequency of HER2+ tumors (11%-58%) compared to the ∼2% population rate as assessed by modern standards, suggesting inclusion of patients without true HER2 positivity, potentially hindering validity of these studies^16^. In other GI malignancies, especially gastroesophageal cancer, HER2 is a strong predictive biomarker^17,18^.

Several immune-based treatments have been studied for PDAC, including peptide vaccine, recombinant vaccine, and irradiated whole tumor cell vaccines^19^, adoptive cell transfer, CAR-T therapy and checkpoint inhibitors^20^. These interventions have had minimal efficacy^21^. For example, a recent study of 50 treatment-naïve patients with advanced PDAC treated with combination gemcitabine, nab-paclitaxel, and nivolumab exhibited a median overall survival of only 9.9 months^22^. Several ongoing phase I or II studies using checkpoint inhibition combined with chemotherapy, radiotherapy, or vaccine therapy such as GVAX and personalized mRNA vaccines, are underway in PDAC^21,23,24^. In gastroesophageal cancer, combination trastuzumab and pembrolizumab recently received FDA approval for first-line treatment based on a response rate of 74%, including an 11% complete response rate^25^. However, in PDAC, combination anti-HER2 therapy with immune checkpoint inhibition has not been reported.

Organoid modeling is an in vitro method that allows dissociated primary tissue, including tumors, to be propagated in three-dimensional tissue culture onto a physical scaffold (matrix) ^26–28^. Patient-derived organoids can be generated from tumor biopsies by cultivation in submerged ECM, such as Matrigel/BME, or can be grown in air-liquid interface (ALI) culture by embedding freshly minced tissue in a collagen bed^29^. Gene expression profiling of organoids can predict tumor responses to therapy^30^ and in vitro testing of drug panels can permit personalized drug screening^31^. In this study, organoids were generated from a patient’s tumor and suggested response to anti-HER2 therapy.

We report a patient with PDAC and HER2 overexpression whom we treated with anti-HER2, immunotherapy, and radiation (RT) combination treatment. In our patient, multiple lines of evidence indicated a high-copy HER2 amplification, raising speculation that her tumor might be driven by HER2 over-activity and thus sensitive to HER2 inhibition. The choice of an anti-HER2 treatment component was motivated by prior studies showing high responses in HER2+ gastric cancer^32^, and organoid modeling experiments predicting sensitivity to anti-HER2 inhibition. On progression of disease, recent advances in anti-HER2 therapy combined with RT and immunotherapy^25^ motivated the use of immune-based treatments, which included checkpoint inhibition and vaccine therapy. Following combined therapy, the patient achieved a durable complete clinical response. Overall, these findings suggest combining anti-HER2 therapy with RT and immunotherapy may be effective for the PDAC patient population with HER2 overexpression.

### Case Report

A previously healthy woman in her 50s presented with an elevated CA-125 level, which had been ordered by her primary care physician as part of a quarterly tumor marker screening panel. On questioning, the patient endorsed abdominal discomfort. An initial pelvic ultrasound observed a large adnexal mass concerning for suspected ovarian cancer. MRI of the abdomen and pelvis identified a 6.3 cm left cystic ovarian mass, a 3.2 cm right ovarian mass, a 3.9 cm pancreatic mass, and at least two liver lesions. In August of 2017, she underwent a hysterectomy, bilateral salpingo-oophorectomy, and infragastric omentectomy. Surgical pathology demonstrated a 10-cm conglomerative omental metastatic mass and bilateral ovarian involvement of a moderately differentiated mucinous adenocarcinoma. Immunohistochemistry staining was notable for CD7 positive, CD20 weak, PAX8 negative, WT1 negative, and DPC4 absence. Histologically the tumor cells contained a pale cytoplasm with mucinous features, strongly suggestive of a metastasis from a pancreaticobiliary primary site rather than an ovarian primary. CT imaging three weeks following surgery identified both an unresected distal pancreatic body mass and concomitant metastatic disease in the liver and hemidiaphragm. Germline genetics evaluation with the Invitae Multi-Cancer Panel identified no pathogenic mutations. Tumor mutational profiling results were pending at the time that systemic therapy was initiated.

In September 2017, the patient began treatment with gemcitabine and protein-bound paclitaxel, combined with indoximod, an investigational immunometabolic agent targeting the IDO pathway, as part of a phase I/II clinical trial (Figure 1)^33,34^. The patient responded well to therapy initially. Pre-treatment CA 19-9 was 17,784 U/mL. Following 10 months of therapy, CA 19-9 reached a nadir of 36 u/mL and a clinical partial response was observed. At that time, her disease burden consisted of a 1.2 cm pancreatic tail mass, a 1.1 cm lesion in the liver, and subcentimeter nodules in the lungs, liver, and peritoneum. Tumor profiling results from her initial resection were then obtained which indicated HER2 overexpression and pathogenic DNA mutations (Table 1). Four months later, her disease burden remained stable except for a single peritoneal lesion in the hepatorenal recess, which had grown to 3.1 cm. Given that the metastasis appeared isolated, and that surgery was considered low risk, the hepatorenal recess lesion was resected in November of 2018. Consistent with her initial resection, the peritoneal lesion exhibited HER2 amplification (Table 1). The tissue showed intact expression of mismatch repair proteins. She then received trastuzumab and pertuzumab on the TAPUR trial^35^.

**Table 1:** Molecular Profiling. Molecular profiling results for the patient’s specimens and derived organoids. HER2 amplifications were detected in all tissues assessed. *Pathogenic Missense* and *Structural Variants* were derived from NGS analysis, while variants of uncertain significance are not listed. The mutations detected in the patient’s surgical specimens were also detected in the organoid. Tempus copy number variation data were provided for intent of research use only. TMB = tumor mutation burden (based the number of non-synonymous mutations per megabase); PD-L1 expression was assessed using the IHC 22C3 assay. Mutation values are presented in minor allele frequency (%). NR = not reported.

| Biomarker Type | Collection Date | August 2017 | August 2017 | November 2018 | November 2018 | March 2020 | March 2020 |
| --- | --- | --- | --- | --- | --- | --- | --- |
|  | Tissue Type | Surgical specimen | Surgical specimen | Surgical specimen | Organoid | Biopsy specimen | Biopsy specimen |
|  | Sequencing Assay | Tempus xT | Tempus xE | Tempus xE | STAMP | Tempus xT | Tempus xE |
| Pathogenic Missense | TP53 R342* | 56.8 | 41.4 | 67.7 | 99.8 | 21.3 | 24.7 |
|  | KRAS G12D | 57.7 | 35.3 | 37.3 | 54.7 | 9.6 | 19.4 |
|  | NF1 R1362* | 4.56 | NR | 2.2 | 6.4 | NR | NR |
| Structural Variants | HER2 | >=20 copies | >=20 copies | >=20 copies | 22.8 copies | >=20 copies | >=20 copies |
|  | TOP2A | >=20 copies | 19 copies | 12 copies | NR | NR | 9 copies |
|  | CDK12 | NR | NR | >=20 copies | NR | >=20 copies | >=20 copies |
|  | MYC | amplification | amplification | NR | NR | NR | NR |
|  | FLT3 | amplification | NR | NR | NR | NR | NR |
|  | MITF | amplification | NR | NR | NR | NR | NR |
|  | NF1 | amplification | NR | NR | NR | NR | NR |
|  | PTP4A3 | NR | amplification | NR | NR | NR | NR |
|  | SMAD4 | deletion | deletion | NR | 0.46 copies | NR | NR |
|  | RARA | NR | NR | NR | NR | NR | 9 copies |
|  | TP53 | loss of heterozygosity | loss of heterozygosity | loss of heterozygosity | NR | NR | NR |
| Immune Biomarker | PD-L1 | positive (2%) | not performed | negative (<1%) | not performed | negative (<1%) | not performed |
|  | TMB | 0.85 | 0.8 | 2.1 | not performed | 3.2 | 0.4 |
| HER2 | IHC (Stanford) | Not performed |  | 3+ | 3+ | 3+ |  |
|  | RNA Sequencing (Tempus) | 95+% |  | 99+% | not performed | 99.7+% |  |
|  | Mass Spectrometry (mProbe) | 1870 attomol/microgram |  | 5895 attomol/microgram | 5048 attomol/microgram | quantity not sufficient |  |

**Figure 1:**
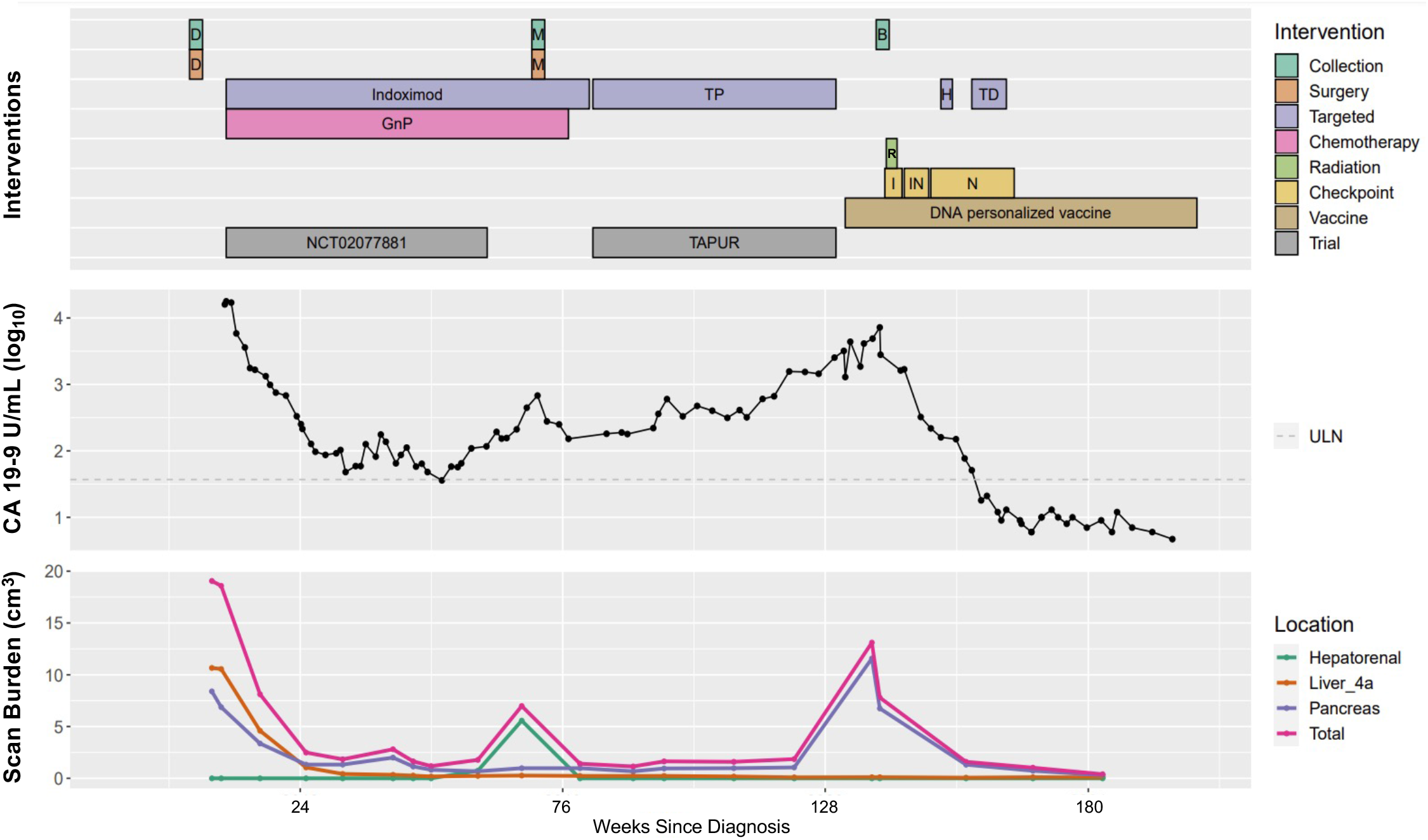
Clinical Timeline. This clinical timeline depicts the interventions delivered and the tumor burden as assessed by CA 19-9 tumor marker and CT scan. Scan burden was measured by approximating the volume of each tumor lesion using the ellipsoid sphere equation, 4/3 * π * A * B * C, where A, B, and C are the lengths of the three semi-axes (radii) of the ellipsoid. Lesions larger than 2 cm at any timepoint are graphed, as well as the total sum of the volumes of each of the 9 lesions present at any time during the scan. D = debulking. M = Metastasectomy. B = Biopsy. H = hydroxychloroquine. TD = trastuzumab deruxtecan. GnP = gemcitabine and nab-paclitaxel. I = ipilimumab. IN = ipilimumab and nivolumab. N = nivolumab. R = Radiation. ULN = upper limit of normal.

On this dual anti-HER2 therapy, the patient’s disease remained stable for 9 months but due to rising CA 19-9 levels she opted to pursue investigational vaccine therapy while continuing trastuzumab and pertuzumab off-trial (Figure 1). In January of 2020, the patient initiated treatment with an investigational neoantigen recombinant DNA vaccine being studied in a phase I trial sponsored by Washington University School of Medicine^36,37^, but as the trial was fully enrolled, the vaccine was provided for compassionate use off-trial. In March 2020, a PET scan showed interval growth of the primary pancreatic lesion in addition to two slow growing ∼1 cm lung lesions in the setting of otherwise stable disease. Multi-disciplinary review among collaborators brought together by the Canopy Cancer Collective pancreatic cancer learning network of recent data suggesting efficacy of concurrent checkpoint inhibition with radiation^38^ and oligometastatic disease^39^ influenced the decision to initiate stereotactic radiation (SBRT) for her pancreatic tumor (40 Gy in 5 fractions) and to the 2 PET avid foci in her bilateral lungs suspected to be metastatic (25 Gy in 1 fraction to LUL and 40 Gy in 4 fractions to the RLL). Imaging and CA 19-9 were consistent with a partial response to treatment. Following completion of radiotherapy, the patient commenced treatment with ipilimumab and shortly thereafter in combination with nivolumab, however the patient discontinued checkpoint inhibition therapy after approximately 6 months due to signs of pneumonitis and acute kidney injury. She was briefly prescribed hydroxychloroquine, but this was discontinued due to nausea and indigestion. Off-label trastuzumab deruxtecan was then added in July 2020 to her ongoing combination immunotherapy with nivolumab and the monthly personalized vaccine. Her CA 19-9 decreased below the upper limit of normal in August 2020. From October 2020, further targeted therapy was held, and only monthly personalized vaccine therapy was continued. By 2021, following approximately four months of combination of targeted anti-HER2 treatment and immunotherapy, imaging showed no evidence of recurrent or metastatic disease. In April 2021, her CA 19-9 measured 4.7, her nadir throughout treatment. As of October 2021, the patient has remained without evidence of disease, asymptomatic and active.

### Molecular Analyses & Organoid Profiling

First-line gemcitabine-based treatment was initiated prior to availability of tumor mutational profiling data, which subsequently showed a *KRAS* G12D mutation, HER2 amplification, PD-L1 positivity (2% of tumor cells), and other abnormalities (Table 1). Thereafter, molecular profiling of the patient’s longitudinal tumor samples and CLIA-grade drug testing of the tissue-derived organoid informed the selection for the targeted and immunooncology therapies she received.

Upon development of a new site of disease in the hepatorenal recess in November 2018, and prior to initiation of therapy, the tissue was biopsied and subjected to whole exome sequencing, RNA sequencing, and organoid generation (Figure 2). Organoids were generated using the air-liquid interface (ALI) culture method^29^ and passaged and maintained using conventional submerged medium culture methods in Cultrex^®^ Reduced Growth Factor Basement Membrane Matrix, Type 2 (BME-2) as described^30^ for subsequent characterization and drug testing. Histological characterization of the generated organoids correlated with original tumor tissue, and the organoid line expressed HER2, assessed by IHC. The whole exome and RNA sequencing results of the organoid matched those from the original surgical sample and the high-copy HER2 amplification was preserved (Table 1). The magnitude of organoid HER2 amplification was substantial, reported as more than 20 copies. Following this molecular validation, the organoid sample was sent to several collaborating institutions for further molecular analysis (Figure 3).

**Figure 2:**
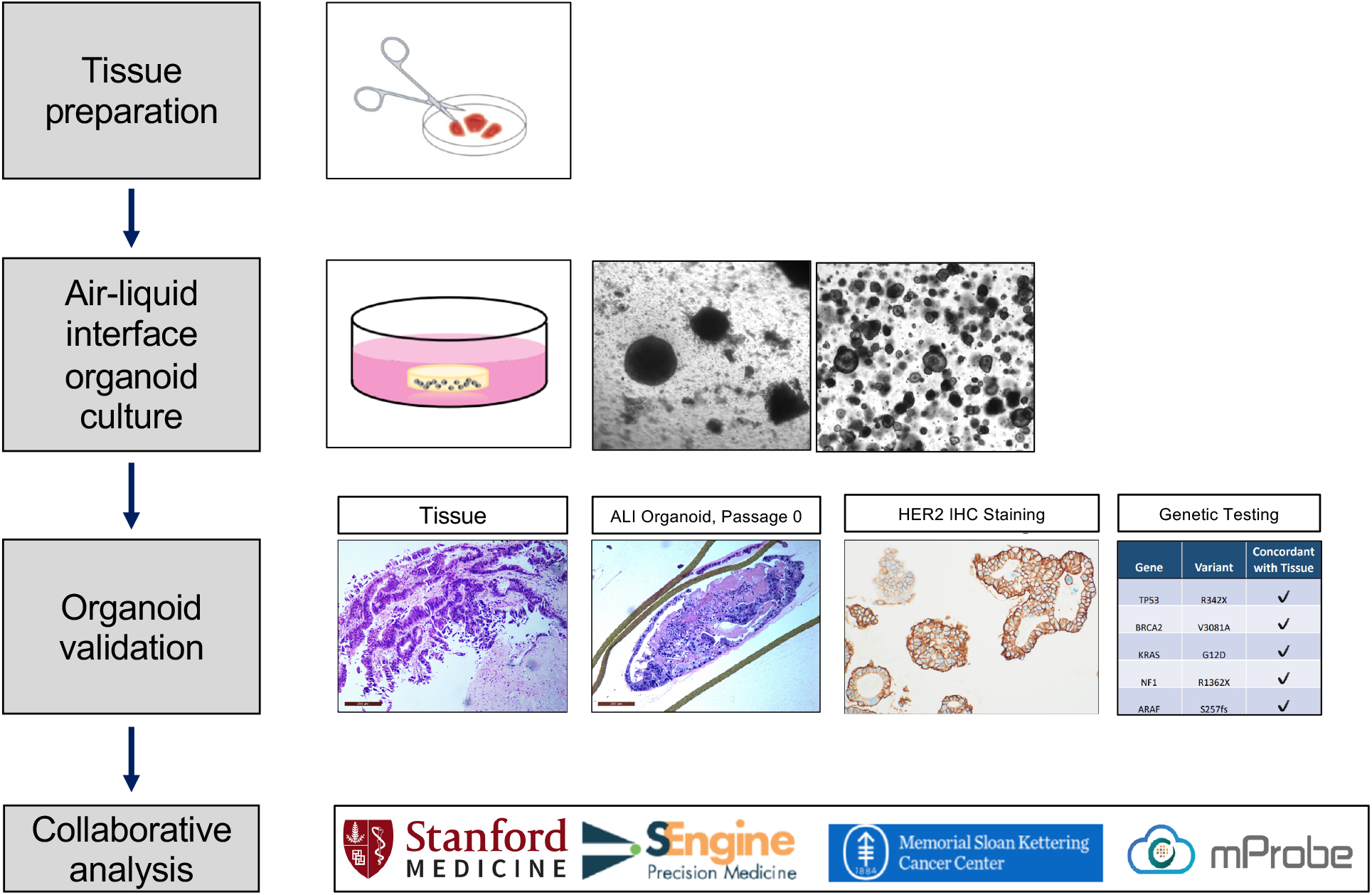
Organoid Generation & Validation. Fresh tumor specimens were minced into small tissue fragments, embedded in a collagen scaffold matrix within an inner transwell, and cultured with direct air exposure above and tissue culture below, contained in an outer dish. Air-liquid interface (ALI) organoids were generated and expanded before being converted into submerged extracellular matrix (BME-2) cultures grown within small domes of matrix beneath tissue culture medium. Organoid validation experiments indicated that the organoid matched the original tissue by histology, had HER2 over-expression, and had a genetic mutations profile concordant with the original tissue. Following confirmation of fidelity, the organoids were distributed to several collaborators for additional study as submerged BME-2 organoids.

**Figure 3:**
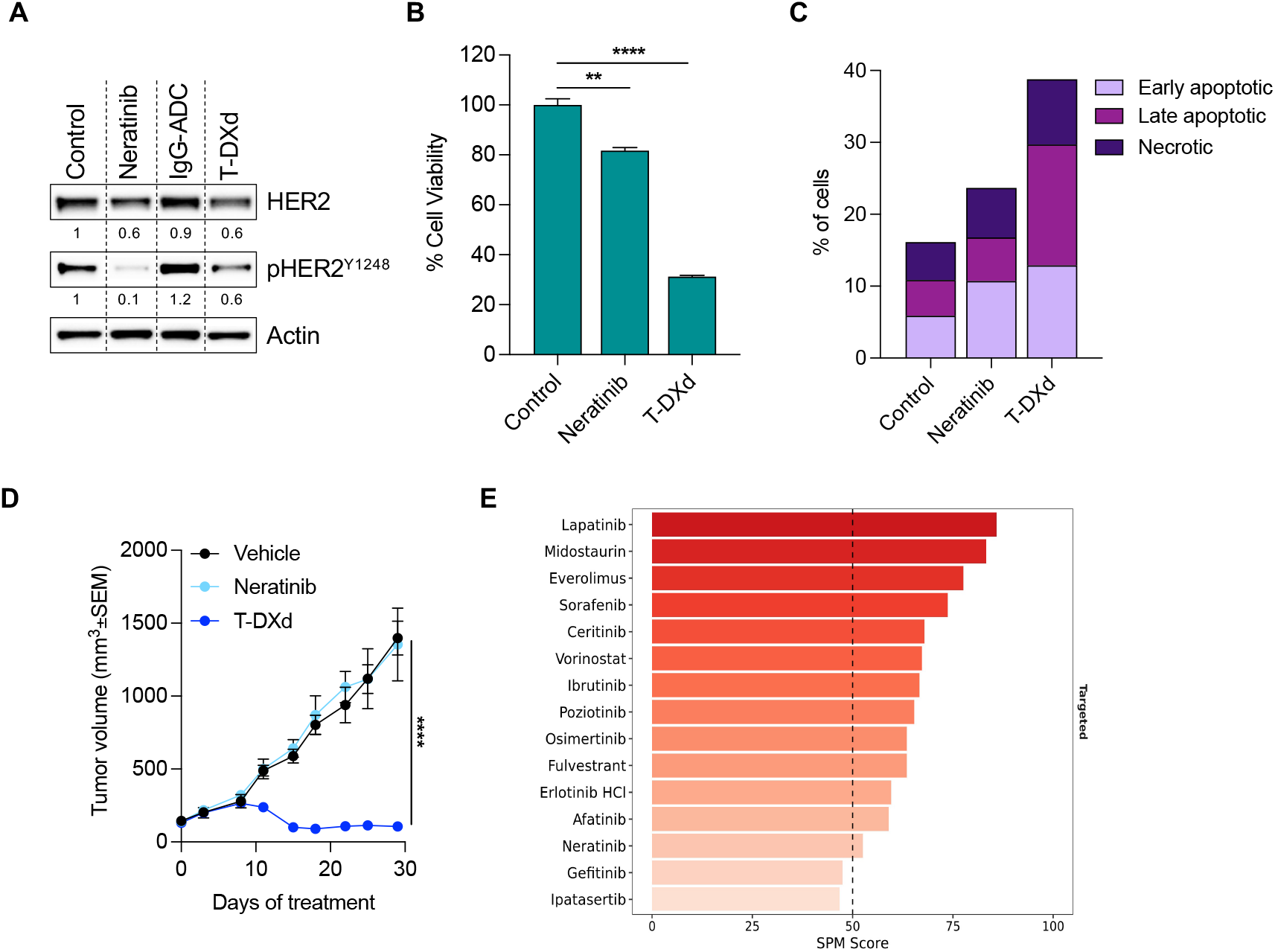
Organoid Analyses. (A) Western blot analysis of patient-derived pancreatic organoids incubated with neratinib (100 nM) or T-DXd (25 μg/mL) or DMSO as control for 6 days. Actin was used as loading control. The numbers represent the amount of total and phospho-HER2 normalized on Actin and relative to Control. (B) Patient-derived pancreatic organoids were incubated with neratinib (100 nM) or T-DXd (25 μg/mL) or DMSO as control for 6 days. Cell viability was assessed by Cell Titer Glo and shown as percentage relative to control ± SEM (n=3). Statistical analyses were performed using t-test (**, P≤ 0.01; ****, P≤ 0.0001). (C) Patient-derived pancreatic organoids were incubated with neratinib (100 nM) or T-DXd (25 μg/mL) or DMSO as control for 6 days. Annexin V staining was measured by flow cytometry and the percentage of early apoptotic, late apoptotic and necrotic cells was shown as stacked bar graph. (D) *In vivo* efficacy study of a ERBB2-amplified pancreatic PDX treated with neratinib (20 mg/kg, orally every day, 5 days a week) or T-DXd (10 mg/kg, i.v. once every 3 weeks). Measurements show average tumor volumes ± SEM, n=5 mice per group. Comparisons between Vehicle and T-DXd groups were performed using two-way ANOVA test (****, P≤ 0.0001 at the indicated time point). E. Top scoring therapeutics from in vitro drug sensitivity testing of organoids as performed using the PARIS^®^ test by SEngine.

CLIA-grade organoid drug sensitivity testing by SEngine demonstrated that anti-HER2 therapy had the highest predicted potency of 39 tested drugs (Supplementary Materials). Organoid drug testing by Memorial Sloan Kettering Cancer Center (MSKCC) also demonstrated sensitivity to anti-HER2 therapy: following a 6-day treatment to the anti-HER2 antibody drug conjugate trastuzumab deruxtecan (T-DxD), approximately 70% of cells died, an effect driven mostly by apoptotic cell death, whereas nearly 100% of cells exposed only to vehicle control remained viable (Figure 3A-C and Supplementary Materials). Xenograft transplantation into immunodeficient mice was attempted for this organoid sample and was not successful; however, an *in vivo* xenograft using organoid tissue from a different patient with PDAC harboring a 4-fold *HER2* amplification and *KRAS* G12D mutation was viable and showed marked reduction in tumor volume after exposure to T-Dxd compared to control (Figure 3D). Lastly, quantitative HER2 expression profiling of organoids using mass spectrometry conducted by mProbe demonstrated a markedly elevated HER2 expression level, above the threshold of predictive sensitivity to anti-HER2 directed therapy^40^ (Table 1). Based on these findings the patient was enrolled on a trial using anti-HER2 directed therapy using trastuzumab and pertuzumab, and her disease remained stable for approximately nine months.

Subsequent follow-up discovered local recurrence in the pancreas as well as clinically diagnosed oligometastatic pulmonary metastases. In January 2020, she began monthly treatments with a personalized DNA vaccine^36^ (Figure 4). A list of the neoantigens incorporated into the vaccine, the mutant and wildtype amino acid sequences, and predicted binding is included in Supplementary Table 2. In March of 2020, SBRT, using 40 Gy over 5 fractions, was delivered. The patient experienced a rapid molecular and radiographic response to this combined radio-immunotherapy. During the fiducial placement for radiation, the pancreatic body primary lesion was biopsied, from which attempted organoid generation was not successful, possibly due to poor tumor viability on-treatment; however, mutation profiling with Next Generation Sequencing (NGS) and RNA sequencing by Tempus on that tissue confirmed continued high-copy HER2 amplification with a > 99.7% rank among the PDAC population assessed by Tempus. This motivated incorporation of continued anti-HER2 directed therapy using trastuzumab deruxtecan (T-DXd), consisting of the monoclonal antibody trastuzumab linked to the topoisomerase inhibitor deruxtecan. Her response continued to deepen on multi-modal therapy, achieving a molecular and radiographic complete response.

**Figure 4:**
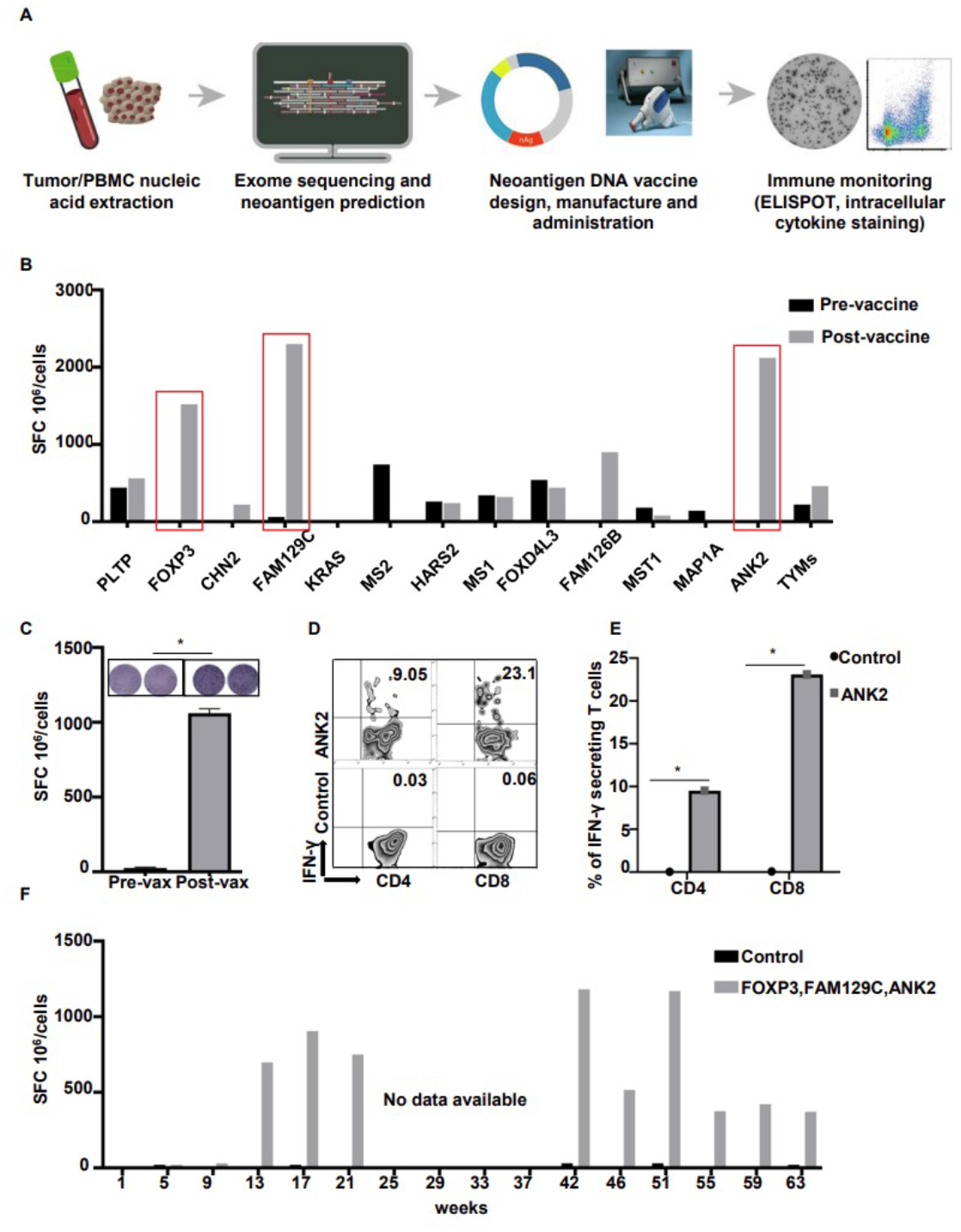
Neoantigen DNA vaccine induces CD4 and CD8 neoantigen-specific T cell responses. (A) Schematic outlining the design, manufacture, administration, and immune monitoring of the neoantigen DNA vaccine. DNA was extracted from both tumor tissue and patient PBMC, while RNA was extracted from tumor tissue only. Tumor/normal exome sequencing was performed to identify somatic genetic alterations. Tumor RNA sequencing was performed to confirm expression of the genetic alterations. The pVAC-Seq suite of software tools was used to identify and prioritize candidate neoantigens. The neoantigen DNA vaccine was designed and manufactured in an academic GMP facility at WUSM. The neoantigen DNA vaccine was administered using an electroporation device. ELISPOT and intracellular cytokine staining were performed to assess the response to vaccination. (B) PBMC obtained before and after vaccination (week 17) were stimulated in vitro for 12 days with peptides corresponding to the indicated neoantigens followed by IFNγ ELISPOT assay. Vaccination induced a strong response to neoantigens FOXP3, FAM129C, and ANK2. (C) ELISPOT response to neoantigen ANK2 before and after vaccination. (D, E) Intracellular cytokine staining demonstrates that ANK2-specific CD4 and CD8 T cell responses were induced. (F) The response to FOXP3, FAM129C, and ANK2 persists over time. PBMC from the indicated time points were stimulated in vitro for 12 days with peptides corresponding to the neoantigens included in the neoantigen DNA vaccine followed by IFNγ ELISPOT assay. The bars indicate the average response to FOXP3, FAM129C, and ANK2. None of the other neoantigens induced a consistent response over time. Nonspecific background counts, assessed by incubating cells without peptide during the ELISPOT assay, were subtracted. Cells stimulated without peptide during the 12-day culture are indicated as a negative control (Control)..

To assess whether the dramatic responses achieved were at least partially attributable to the vaccine, functional studies of immune-driven tumor suppression were performed, which demonstrated that the neoantigen DNA vaccine induced CD4 and CD8 neoantigen-specific T cell responses (Figure 4). IFNγ ELISpot assay performed after in vitro culture of PBMCs collected pre- and post-vaccination (week 17) with pooled neoantigens indicated that the neoantigen DNA vaccine induced robust T cell responses against three neoantigens FOXP3 (p.A349T), FAM129C (p.G520R), and ANK2 (p.R2714H) (Figs 4B, C, and F). Further study by intracellular cytokine staining demonstrated that ANK2-specific CD4 (9.05%) and CD8 (23.1%) T cell responses were induced after stimulation of PBMC with ANK2 (Figs. 1D and 1E). The response to FOXP3, FAM129C, and ANK2 persisted over time (Fig. 1F). Of note, the bars in Fig. 1F indicate the average response to FOXP3, FAM129C, and ANK2. None of the other antigens induced a consistent response over time.

The patient’s blood samples continue to be assessed for circulating disease and she remains in clinical and molecular remission.

## Discussion

This report highlights the case of a patient with metastatic pancreatic cancer and HER2-amplification, who achieved a complete response following multiple lines of targeted and immune-based therapies. Initially, she underwent surgical debulking for suspected ovarian cancer, whereas this procedure is generally reserved for symptomatic or chemo-refractory disease for PDAC. Subsequently, she underwent gemcitabine-based treatment, anti-HER2 therapy, and then third-line combination therapy with an HER2-directed antibody conjugate, radiation, checkpoint, and personalized vaccine therapy. Remarkably, following combination therapy, the patient has achieved a deep and durable remission.

Complete responses are rarely observed in PDAC, representing 0.2% (1 of 431) of patients who received first-line FOLFIRINOX on trial^41^, 0.6% (1 of 171) of patients treated with first-line gemcitabine and nab-paclitaxel on trial^42^, and 0.0% (0 of 212) of patients who received nano-liposomal irinotecan following progression on gemcitabine-based therapy^43^. In a trial of patients with germline BRCA deficiency, 2% (2 of 92) of PDAC patients achieved a complete response on olaparib^44^. Thus, the observation that a complete and durable response was achieved in the third line setting prompts speculation that the patient’s disease biology, treatment regimens, or combination may be explanatory.

The patient’s tumor was notable for HER2 amplification. Multiple lines of evidence resulting from our organoid studies indicated potent sensitivity to anti-HER2 directed therapy. Her disease stability of dual HER2-directed therapy, lasting 9 months, compared to a median expected progression free survival of 4.4 months in the second-line setting^45^, and deep remission achieved on trastuzumab deruxtecan were coincident, suggesting a mechanistic underpinning. As one explanation, this patient had an exceptionally high HER2 copy number state, perhaps suggesting a component of oncogene addiction driven by a quantitative relationship between HER2 amplification and response in PDAC. Accordingly, prior literature in gastric cancer^40^, breast cancer with trastuzumab^46^ and trastuzumab deruxtecan^47^ support a positive correlation between HER2 gene copy number and response to anti-HER2 therapy. Conversely, loss of HER2 expression has been observed as a mechanism of resistance following anti-HER2 therapy in gastric cancer^48,49^. Notably, molecular profiling of the patient’s relapsed tumor showed persistent high-copy HER2 amplification following trastuzumab and pertuzumab which informed subsequent anti-HER2 therapy.

The deepest measured responses achieved, as assessed by a logarithmic drop in CA 19-9 and reduction in volumetric tumor burden, occurred at the initiation of therapy, and during radiation and immunotherapy. While it is possible that the response to radiation and immunotherapy would have been sustained and led to a complete response on their own, anti-HER2 therapy appeared to contribute to and may have potentiated the effect of combined therapy. Of note, a co-mutation in KRAS may predict lack of response to HER2 directed therapy in HER2 amplified colorectal cancer^50^, and gastric cancer^51^ although this may not be the case in pancreatic cancer, and it is conceivable that the magnitude of amplification in our patient led to a dependence and oncogenic addition to the HER2 pathway.

Multi-disciplinary collaboration informed a multi-modality treatment strategy, combining surgery, radiation, checkpoint inhibition, personalized vaccine, and anti-HER2 drug antibody conjugate. These treatments were given in a partially concurrent manner to leverage additive benefits observed in pre-clinical and early clinical settings and were staggered to mitigate toxicity. Trastuzumab deruxtecan combined with immunotherapy potentiates a strong immune response and acts synergistically with anti-PD-1 antibody treatment to prolong survival in mice^52^. The combination of radiation with immunotherapy may have an abscopal effect to induce immune response to neoantigens, a theory being studied across multiple cancer types ^53^, showing activity in colorectal cancer^54^ and anecdotal evidence in pancreatic cancer^55^. Combination vaccine therapy with checkpoint inhibition indicates that PD-1 blockade increases CD8 positive effector T cells and prolongs mouse survival compared to vaccine therapy alone^56^. Another preclinical study combining a neoantigen vaccine with checkpoint inhibition in mice demonstrated prolonged survival compared to vaccine or checkpoint therapy alone^57^.

While our organoid modeling was effective for understanding tumor cell-autonomous HER2 response, it did not allow evaluation of the ALI organoid tumor immune cellular compartment, which does not persist long-term; immunotherapy was not a therapeutic consideration at the time of the initial biopsy and organoid generation. However, acute evaluation of checkpoint inhibition within ALI tumor cultures is feasible within the first several weeks of culture and can be considered for future cases^29,58^ Also of note, the initial therapy with the immunomodulator agent indoximod, which elicited a partial response, may have primed the subsequent response to checkpoint inhibition. While the phase II trial of the indoximod-containing regimen received by the patient failed to meet its primary endpoint^59^, exploratory analyses showed that responders had increased intra-tumoral CD8 density compared to non-responders (p=0.30). The patient was prescribed hydroxychloroquine based on data suggesting an important role of autophagy in regulating MHC-I mediated immunogenicity in PDAC^60^, but this was stopped due to intolerance. Lastly, functional assessment of the patient’s personalized DNA neoepitope vaccine demonstrated that her T cells were activated against cancer-specific epitopes, thus providing evidence that the vaccine is now contributing at least partially to response and maintenance of remission.

An overall survival advantage for biomarker-directed therapy in PDAC has yet to be demonstrated in a prospective, randomized trial. Nevertheless, several findings suggest that a subset of PDAC patients with targetable alterations are likely to achieve benefit from biomarker-matched therapy. For example, a retrospective, non-randomized study involving PDAC patients eligible for biomarker-matched therapy based on having microsatellite instability, DNA repair gene mutations, or other mutations, had impressive survival as compared to historical averages of unselected PDAC patients^61^. Additionally, long duration of response to olaparib has been observed in patients with germline BRCA mutations^44^. Among the 5-10% of PDAC patients who lack RAS mutations, an enrichment of HER2 mutations, other targetable gene mutations, and microsatellite instability are observed^62^. NCCN guidelines currently recommend that PDAC patients with advanced disease undergo tumor panel gene profiling, including HER2 mutations, and mismatch repair assessment, but do not explicitly recommend HER2 IHC or amplification testing. Anecdotal evidence of exceptional cases may help identify strategies that correlate patient attributes or tumor biomarkers with predictive value in improving outcomes. Collaborative, multi-disciplinary trials combining radiotherapy, immunotherapy, and targeted therapy, may exploit tumor vulnerabilities to guide PDAC precision therapy as exemplified by the current patient.

## Data Availability

All data produced in the present work are contained in the manuscript, data are available upon reasonable request for purpose of peer review.

## Acknowledgements

The investigators are indebted to our patient for the permission to publish her clinical story. Informed consent to publish information and/or images from the patient. We appreciate guidance from Maurizio Scaltriti for several organoid analyses. Drs. Sunil Hingorani, Andrew Hendifar, and Ted Hong provided valuable clinical insight. We are grateful to Kevin White, Greg Call, Nike Beaubier, and Amber Solari, at Tempus for sharing HER2 distribution data for research use. We are grateful for support from the NIH (U01CA217851, U54CA224081, R01CA2515143), Seed Grant from the Stanford Cancer Institute, NIH NCI Award (U01CA248235), and V Scholar Award from the V Foundation for Cancer Research (V2018-007).

## Figures

See PowerPoint

## Supplementary Materials

### Organoid Generation

Core biopsy samples were transported to the lab in HypoThermosol ^®^ FRS preservative solution (StemCell Technologies, #07935) on ice. As described previously, the biopsy sample was minced, resuspended in collagen type 1 matrix and transferred into an inner transwell containing a base layer of collagen type 1 matrix (Fujifilm Wako Chemicals, Cellmatrix Type 1-A, #631-00651)^1^. Organoids were generated and maintained in culture using WENR media as described in Neal, et al, 2018. Upon ALI organoid generation, collagen was dissociated using collagenase and organoids dissociated using TrypLE Express (Gibco). Dissociated organoids were resuspended in Cultrex^®^ Reduced Growth Factor Basement Membrane Matrix, Type 2 (BME-2) (R&D Systems) for submerged organoid culture.

**Supplementary Figure 1:**
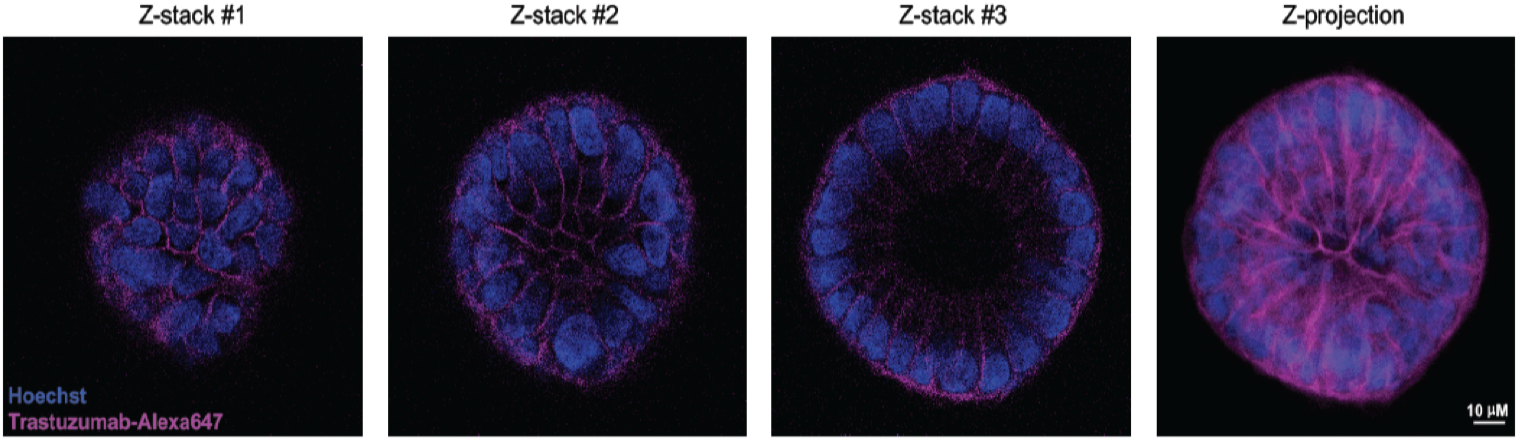
Confocal Immunofluorescence. Confocal immunofluorescence images from submerged organoids collected 1 hour post-trastuzumab treatment demonstrate the 3D organoid colony. This image confirms trastuzumab localization to cellular membranes in HER-2 expressing organoids.

### MSK Studies

#### Western Blot

Organoids were plated into 6-well plates and treated with DMSO, neratinib 100nM or T-DXd 25 μg/mL for 6 days. Corning^®^ Cell Recovery Solution was used to recover organoids cultured on Corning^®^ Matrigel^®^ according to manufacturer’s instructions. Total protein lysates (20 μg) were extracted using RIPA buffer and separated on SDS-PAGE gels (NuPAGE 4–12% Bis-Tris Protein Gels, Invitrogen) according to standard methods. Membranes were probed using the following antibodies: anti-total HER2 Rabbit mAb (29D8, Cell Signaling Technology #2165), anti-phospho-HER2 (Tyr1248, Cell Signaling Technology #2247) and anti-β-Actin 13E5 (Cell Signaling Technology #4970). ImageJ was used to quantify western blot band intensity.

#### Cell Viability

The CellTiter-Glo luminescent cell viability assay was performed on each sample according to the manufacturer’s instructions (Promega, G7570). Briefly, organoids were plated in triplicate onto 96-well plates. After an incubation for 24 h, organoids were treated with DMSO, neratinib 100nM or T-DXd 25 μg/mL for 6 days. Organoids were then incubated for 1 hour at 37 degrees Celsius with CellTiter-Glo reagent, and luminescence was measured using a 96-well plate reader. Background luminescence was measured in medium without organoids and subtracted from experimental values.

#### Flow Cytometry

Organoids were plated in a 12-well plate and cultured in presence of DMSO, neratinib 100nM or T-DXd 25 mg/mL for 6 days. For the apoptosis assessment, cells were collected, resuspended, and analyzed for phosphatidylserine exposure by staining with DAPI and Annexin V APC (550474; BD Bioscience) after incubation with Annexin V binding buffer (556454; BD Pharmingen) according to the manufacturer’s instructions. A minimum of 10.000 cells were acquired using FACS CANTO II instrument (BD, Heidelberg, Germany) and data was analyzed using FlowJo v10 software (Tree Star, Ashland, OR).

**Supplementary Figure 2:**
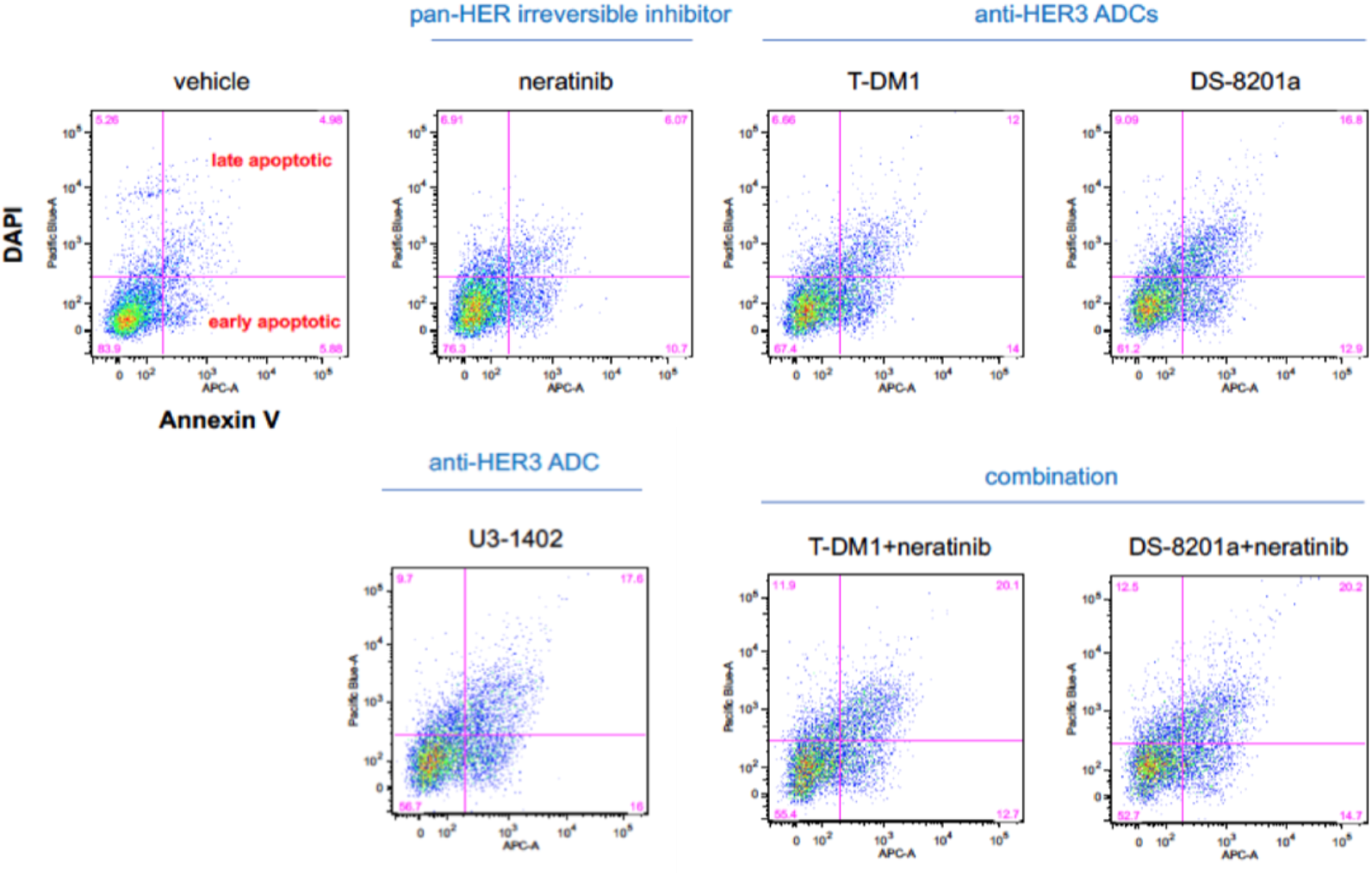
MSK Flow Cytometry Data: FACS analysis of PDAC-35T organoids after 6-days treatment. Neratinib = panHER irreversible inhibitor, T-DM1 = anti-HER2 ADC, DS-8201a = anti-HER2 ADC, U3-1402 = anti-HER3 ADC.

### In Vivo Study

The ERBB2-amplified pancreatic patient-derived xenograft (PDX) was generated as follows: 6-week-old NOD SCID gamma female mice were implanted subcutaneously with specimens freshly collected from a patient at MSK under an MSK-approved IRB bio-specimen protocol #06-107. Tumors developed within 2 to 4 months and were expanded into additional mice by serial transplantation. At this point, the PDX was subjected to high-coverage NGS with the MSK-IMPACT assay. For the efficacy study, treatment was started when tumor volumes reached approximately 150 mm^3^. Xenografts were randomized and dosed with neratinib (20 mg/kg, orally 5 days a week), T-DXd (10 mg/kg, intravenously once every 3 weeks), or vehicle as control (saline, orally 5 days a week). Mice were observed daily throughout the treatment period for signs of toxicity. Tumors were measured twice weekly using calipers, and tumor volume was calculated using the formula length × width2 × 0.52. Body weight was also assessed twice weekly. At the end of each treatment, animals were sacrificed, and tumors were collected for biochemistry and histology analysis. Mice were cared for in accordance with guidelines approved by the MSK Institutional Animal Care and Use Committee and Research Animal Resource Center. Five (5) mice per group were included in the experiment.

### SEngine Studies

SEngine Precision Medicine (SPM) received patient-derived organoids (PDO) from Dr. Calvin Kuo’s laboratory, which were derived from a case of pancreatic adenocarcinoma (PDAC) with peritoneal metastasis. Organoids were expanded for 4 weeks at SPM and plated for the drug sensitivity CLIA-certified assay, PARIS® Test, measuring drug response across a panel of oncology drugs. For each case, drugs are selected based on the cancer type, genomic knowledge, as well as the SPM knowledge base where available, as well as the doctor’s recommendations. The current PARIS test aims to test up to 44 drugs, provided that sufficient material is obtained. At the time this test was performed, the results were ranked with a score from 100 - 1, with drugs that exhibited any degree of sensitivity receiving a score between 100 to 50, generally.

Since 2020, categories have been added to numerical scores and the score has been condensed from 15 to 1. The recent clinical analysis of the scorings system and categories has indicated a strong correlation (∼70%) in both the retrospective and prospective setting^2^. Curated categories that characterize drug responses have been added comprising: *Exceptional, Good, Moderate, Low* and None. Only drugs scoring at least *Low* are presented.

The drug panel for this patient consisted of 39 targeted drugs, each tested at 6 dilution points as specified further below covering the following targets and pathways: EGFR & HER2, PARP, ER, MEK, PI3K/AKT/mTOR, FLT3, BET, BCR-ABL/SRC and VEGFR.

The results of the PARIS® assay indicated lapatinib as a top-scoring drug with a categorization of *Exceptional/Good*, borderline between exceptional to good, with the updated scoring system. Lapatinib, a HER2 small molecule inhibitor, is used in the PARIS^®^ Test as a surrogate for HER2-directed therapies such as trastuzumab. This result was in concordance with the presence of high levels of HER2 amplification and overexpression as detected by genomics and IHC clinical tests. In addition, EGFR targeting poziotinib, ibrutinib, osimertinib and erlotinib all showed *Moderate* response. Other top scoring drugs included midostaurin, a multi-kinase inhibitor developed for FLT3 mutant leukemias. Interestingly, also quizartinib, a FLT3 inhibitor, indicated a *Moderate/Low* response. In addition, a *Good* response was detected to Everolimus, and a *Low* response to the AKT inhibitor, ipatasertib, the latter is typical for this class of drugs that are best used as combination therapies. Of note, despite the presence of a KRAS G12D mutation, this case did not demonstrate sensitivity to MAPK inhibitors used as single agents, such as trametinib or cobimetinib.

Description of the PARIS® Test

The PARIS® Test is a high throughput and high complexity CLIA-certified assay applied to cancer specimens grown with specific media and conditions for each cancer type. The media and procedures have been optimized at SPM to promote selection of tumor cells over stroma and other cellular components, such as lymphocytes. For this case, already established organoids were received by Dr. Calvin Kuo’s laboratory and expanded using standard conditions for PDAC organoids.

#### HTS assay

Briefly, the PARIS® Test assay is carried out in 384-well format and drugs are assayed with a 6 point-drug titration spanning generally a range of concentrations, from Log10e-7.5(M) to Log10e-5(M). Depending on the therapeutic range, certain drugs may be tested within a different set of concentrations. Methods pertaining the high-throughput drug sensitivity assay, including drug combination studies, were as described in previous publications^3,4^.

**Supplementary Table 1:** SEngine Top Response Drugs. Table of top-scoring targeted drugs displayed with corresponding targets, maximum observed serum dose (Cmax), and IC50. Drugs are ranked by SPM score, which is a proprietary ranking metric of drug sensitivity that weighs sensitivity and uniqueness of response obtained through the PARIS® Test, on a scale with values from 100 - 1 ranked from the best to the worst for each patient. High numbers (100 - 50) indicate drugs with some degree of response, while a SPM score below 50 generally indicated low to no response. Rank is of guidance but should not be considered the only metric to select drugs as clinical consideration and genomic evidence should be considered for example.

| Drug | Target | Cmax | Inhib.Cmax | IC50 | SPM | Interpretation |
| --- | --- | --- | --- | --- | --- | --- |
| Lapatinib | EGFR, HER2 | 4.00E-06 | 91 | 9.70E-07 | 85.9 | Exceptional/Good |
| Midostaurin | FLT3, PKCα, PKCβ, PKCγ, Syk, Flk-1, Akt, PKA, c- Kit, FGFR, SRC, PDFRβ, VEGFR1, VEGFR2 | 1.40E-06 | 64.3 | 3.50E-07 | 83.3 | Good |
| Everolimus | mTORC1 | 3.90E-08 | 31.3 | 1.40E-06 | 77.6 | Good |
| Sorafenib | VEGFR, PDGFR, Raf | 2.10E-05 | NA | 2.00E-06 | 73.7 | Good |
| Ceritinib | ALK, IGF-1R, ROS1 | 1.40E-06 | 49.4 | 1.50E-06 | 67.9 | Moderate |
| Vorinostat | histone deacetylase | 2.20E-06 | 58.6 | 1.70E-06 | 67.3 | Moderate |
| Ibrutinib | BTK | 8.70E-07 | 40.9 | 2.20E-06 | 66.7 | Moderate |
| Poziotinib | EGFR, HER2, HER4 | 1.70E-07 | 57.3 | NA | 65.4 | Moderate |
| Fulvestrant | selective estrogen receptor degrader | 2.10E-08 | NA | NA | 63.5 | Moderate/Low |
| Osimertinib | EGFR | 1.30E-07 | 24.3 | 1.50E-06 | 63.5 | Moderate/Low |
| Erlotinib HCl | EGFR, ALK, JAK2 mutant (JAK2V617F) | 5.40E-06 | 77.8 | 7.70E-07 | 59.6 | Moderate |
| Afatinib | EGFR, HER2 | 3.90E-07 | 46.1 | 1.40E-06 | 59 | Low, IC50 is higher than Cmax |
| Neratinib | EGFR, HER1, HER2, HER4 | 2.10E-07 | 33.3 | 6.30E-07 | 52.6 | Low |
| Gefitinib | EGFR | 3.60E-07 | 28.8 | 2.10E-06 | 47.4 | Low |
| Ipatasertib | pan-AKT | NA | NA | NA | 46.8 | Low, partial response 30% |

### Trastuzumab Drug Combination Testing

Subsequently, a drug combination study aiming to identify potential drugs to enhance the effect of trastuzumab was performed as a custom research study. Trastuzumab was titrated onto the PDOs to obtain the IC30 concentration which was determined at 200 ng/ml. This single concentration of trastuzumab was employed as a “sensitizer” against an additional panel of 24 drugs. Results indicated enhanced inhibition of PDO growth with the combination of trastuzumab with the MEK inhibitor cobimetinib. This drug combination testing revealed a “hidden” sensitivity to inhibition of the MAPK pathway, which emerged upon inhibition of the HER2 receptor. This result may be explained by the presence of the KRAS G12D mutation, which generally correlates with a strong response to MEK inhibitors. In addition, the drug combination study showed enhanced growth inhibition by the combination of trastuzumab and adavosertib. The latter targets the cell cycle checkpoint kinase WEE1. The presence of a TP53 stop gain mutation (p.R343) in this specimen may underline the sensitivity of this specimen to adavosertib.

### mProbe Studies

Selected reaction monitoring-mass spectrometry (SRM-MS) of 72 biomarkers of the tissue sections from FFPE blocks was conducted as previously described^5,6^. Briefly, two tissue section sections (10µM each) from FFPE blocks were placed on DIRECTOR slides, deparaffinized, and stained with hematoxylin. Tumor areas were marked by board-certified pathologist which was microdissected and solubilized to tryptic fragments as per manufacturer instructions (Expression Pathology, Rockville, MD). Protein concentrations of the tryptic peptides was calculated using microBCA. Stable heavy isotope-labeled internal standard peptides for 72 biomarkers were added to the solution and injected into the mass spectrometer (TSQ Quantiva, ThermoFisher Scientific, San Jose, CA). On-column injection resulted in 5fmol of isotopically labeled internal standard peptide and 1µg of total tumor protein. Data analysis of the 72 biomarkers was conducted using Pinnacle software (Optys Tech, Boston, MA).

### Personalized Vaccine Therapy

#### Regulatory approval of the neoantigen DNA vaccine

The neoantigen DNA vaccine treatment protocol was approved by the Washington University School of Medicine (WUSM) Institutional Review Board, the Institutional Biosafety Committee, and the Food and Drug Administration. Written informed consent was obtained from the patient.

#### Neoantigen DNA vaccine design and manufacture

DNA from both tumor tissue and peripheral blood mononuclear cells (PBMCs) was extracted using the QIAamp DNA Mini Kit (Qiagen Sciences, Maryland), and RNA was extracted from tumor tissue using the High Pure RNA Paraffin kit (Roche, Indianapolis). DNA and RNA quality and quantity were assessed using a Nanodrop 2000 and a Qubit Fluorometer (Life Technologies, Carlsbad, CA), respectively. Exome sequencing and cDNA-capture sequencing were performed at the McDonnell Genome Institute at WUSM followed by selection and prioritization of candidate neoantigens using the prediction algorithms in the pVAC tools suite of software^7^. The candidate neoantigens were subsequently cloned into a pING plasmid backbone, as previously described^8,9^. In addition, two epitopes derived from mesothelin, a pancreas cancer-associated tumor antigen, mesothelin were included^10–12^. The neoantigen DNA vaccine was manufactured at the Biologic Therapy Core Facility at WUSM. Extensive product release tests were performed to confirm the identity of the plasmid, and the suitability of the plasmid for administration prior to release (Fig 1A).

**Supplementary Table 2:**
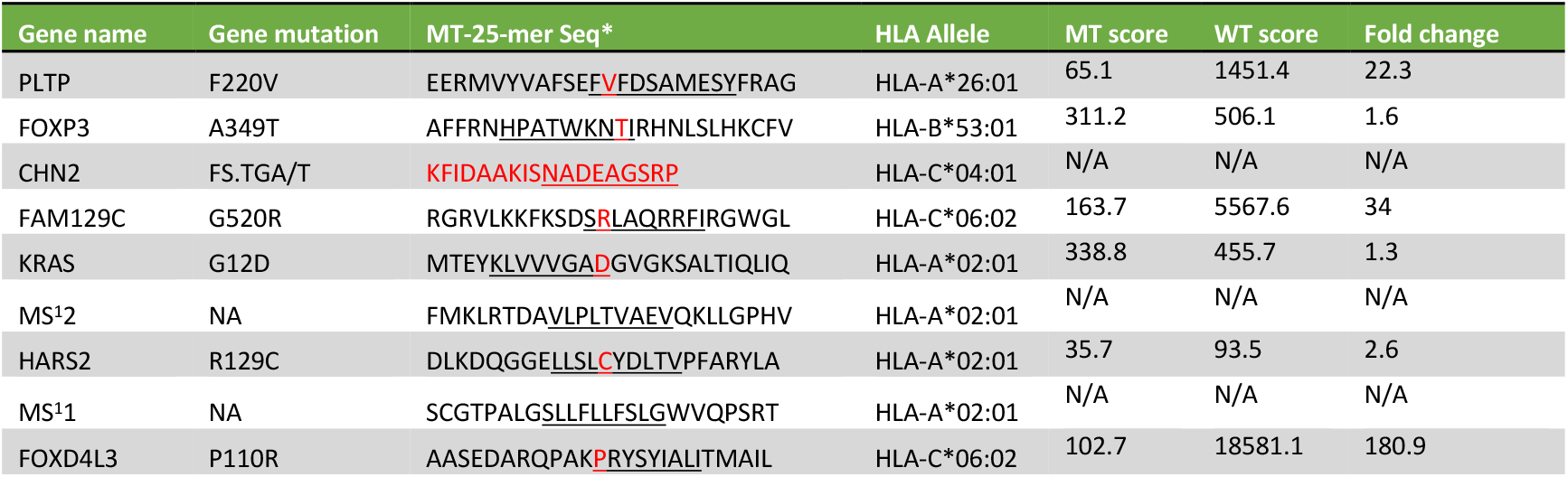
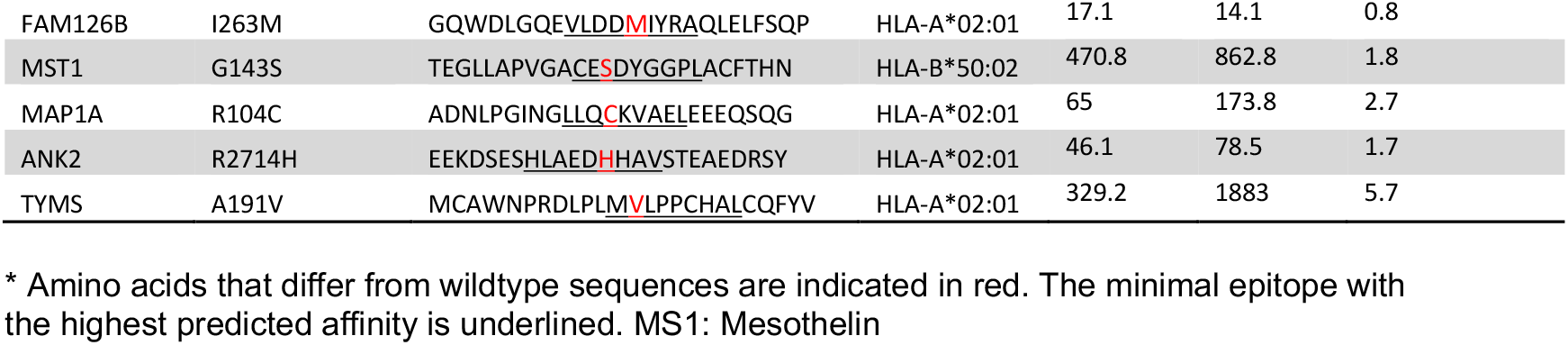
Neoantigens included in the neoantigen DNA vaccine

#### Vaccine administration

The neoantigen DNA vaccine was administered monthly using an integrated electroporation device (TDS-IM system, Ichor Medical Systems, San Diego, CA). At each vaccine time point, the patient received two injections of 2 mg DNA vaccine (4 mg total at each time point). To date, the patient has received 16 months of vaccinations. At each vaccine time point, peripheral blood was drawn and PBMCs were isolated by Ficoll-Paque PLUS (GE Healthcare) density centrifugation and cryopreserved.

#### ELISPOT assay

IFN-γ ELISpot^PLUS^ Kits (Mabtech, Cincinnati, OH) were used according to the manufacturer’s instructions and as detailed below to measure neoantigen-specific immune responses^8,9^. Overlapping synthetic peptides of 15-16 amino acids in length corresponding to the neoantigens included in the neoantigen DNA vaccine were synthesized by LifeTein (Hillsborough, New Jersey, USA). Three peptides overlapping by 11 amino acids were synthesized for each neoantigen included in the vaccine. Synthetic peptides corresponding to the two mesothelin epitopes were also synthesized. Cryopreserved PBMCs were thawed, and typically plated at 2 × 10^5^ per well in duplicate or triplicate followed by 12 days *in vitro* stimulation with pooled peptides. During the 12-day culture, overlapping peptides (2 µM) for two candidate neoantigens were pooled and added to PBMC in the presence of human IL-2 (50 U/mL). After 12 days, lymphocytes were harvested and rested overnight in culture medium without peptides and IL-2. The next day, 10^5^ of the rested cells were co-cultured in the ELISpot plate for 20 h with 10^4^ of irradiated (3000 Rad) autologous PBMCs pulsed with (or without) peptides (5 µM) corresponding to the neoantigens used in the 12-day cultures. The ELISpot plates were scanned and analyzed on an ImmunoSpot Reader (CTL, Shanker Heights, OH).

#### Flow cytometry

The following anti-human monoclonal antibodies (mAb) were used for staining: live/dead AF488 (ThermoFisher Scientific, Waltham, MA), CD4-PerCP-Cy5.5 (clone: RPA-T4), CD8-PE (clone: HIT8a), and IFN-gamma-APC (clone B27). All antibodies were purchased from BD Bioscience (San Jose, CA). Samples were acquired on a flow cytometer (BD Biosciences), and data were analyzed using FlowJo software.

#### Statistics

Data were analyzed using GraphPad Prism 9 software (GraphPad, La Jolla, CA) and presented as mean or the mean ± SEM, where appropriate. The Student *t* test and one-way ANOVA were used to compare between data sets. A *P* < 0.05 was considered statistically significant.

## Notes

### Competing Interest Statement

The authors have declared no competing interest.

### Author Declarations

Ethics committee/IRB of Northwell Health name waived ethical approval for this work. No PHI data are disclosed.

